# Tooth Loss, Oral Health-Related Quality of Life, and Sexual Function in Women

**DOI:** 10.64898/2026.07.09.26357487

**Authors:** Viviane Moura Novaes, Rodolfo Macedo Cruz Pimenta, Caroline Santos Silva, Belarmino Victor Sobrinho Netto, José de Bessa Júnior, Márcio Campos Oliveira

## Abstract

**Objective:** To evaluate the association of tooth loss and oral health-related quality of life (OHRQoL) with sexual function in adult women attending a primary dental care service.

**Methodology:** This cross-sectional study included 99 sexually active women aged 19-66 years who were consecutively recruited from January to October 2023. Tooth loss was quantified by standardized oral examination. OHRQoL was assessed using the Oral Health Impact Profile-14 (OHIP-14), and sexual function was assessed using the Female Sexual Function Index (FSFI). Sexual dysfunction was defined as FSFI <=26.5. Spearman rank correlation was used for bivariate analyses. Multivariable logistic regression was used to evaluate factors associated with sexual dysfunction, including number of missing teeth, OHIP-14 score, age, and relationship status.

**Results:** Tooth loss was present in 83.8% of participants, with a median of 4 missing teeth (interquartile range [IQR], 1-10). Sexual dysfunction was identified in 62.6% of women. FSFI scores were negatively correlated with number of missing teeth (rho = −0.407; p < 0.001), OHIP-14 score (rho = −0.279; p = 0.005), and age (rho = −0.334; p < 0.001). In multivariable logistic regression, OHIP-14 score was independently associated with sexual dysfunction (OR = 1.05; 95% CI, 1.01-1.10; p = 0.015), whereas number of missing teeth was not independently associated after adjustment.

**Conclusion:** Worse OHRQoL was independently associated with sexual dysfunction, whereas tooth loss was associated with lower FSFI scores only in bivariate analysis. These findings are compatible with the hypothesis that the impact of tooth loss on sexual function may be partly explained by oral health-related quality of life, but longitudinal studies are required to test causal and mediational pathways.

## Introduction

Tooth loss remains a major oral health problem and is closely linked to oral health-related quality of life (OHRQoL). Beyond functional limitations related to chewing and speech, missing teeth may affect appearance, self-perception, social confidence, and interpersonal relationships. These psychosocial consequences are especially relevant in settings with high burdens of untreated oral disease and limited access to rehabilitation.

Sexual function is a multidimensional aspect of well-being influenced by biological, psychological, relational, and social determinants. Although oral health and sexual function are usually investigated separately, tooth loss may plausibly affect sexual well-being through body image, self-esteem, social avoidance, and reduced quality of life. This pathway is clinically relevant because oral rehabilitation may improve not only oral function but also broader dimensions of well-being.

Evidence directly linking tooth loss, OHRQoL, and sexual function in women remains limited. Most available studies have focused on general quality of life, older populations, or prosthetic rehabilitation, with fewer data from women attending primary dental care. The aim of this study was to evaluate the association of tooth loss and OHRQoL with sexual function in adult women attending a primary dental care service.

## Material and Methods

Study design and participants. This cross-sectional study was conducted in a primary dental care service between January and October 2023. Reporting followed the Strengthening the Reporting of Observational Studies in Epidemiology (STROBE) recommendations. Women were consecutively recruited if they were adults, sexually active, and free of decompensated chronic disease, defined as uncontrolled diabetes or uncontrolled hypertension. Women undergoing treatment for sexual dysfunction were excluded. All participants provided written informed consent, and the study protocol was approved by the Institutional Review Board of Universidade Estdual de Feira de Santana (approval number 6.199.871, dated July 25, 2023; CAAE, Brazilian ethics platform registration number 58114722.0.0000.0053).

Oral assessment. Participants underwent a standardized oral examination performed by a single trained dentist. Tooth loss was quantified as the number of missing teeth and analyzed as a continuous variable. For descriptive purposes, edentulism was also classified into five categories: no missing teeth; loss of up to 12 posterior teeth; loss of up to 12 teeth including anterior teeth; loss of more than 12 teeth; and complete edentulism. OHRQoL was assessed using the Oral Health Impact Profile-14 (OHIP-14), a validated short-form instrument in which higher scores indicate greater negative impact of oral health on quality of life.

Sexual function. Sexual function was assessed using the Female Sexual Function Index (FSFI), a multidimensional self-report instrument that evaluates desire, arousal, lubrication, orgasm, satisfaction, and pain. Total scores range from 2 to 36, with higher scores indicating better sexual function. Sexual dysfunction was defined as FSFI <=26.5, according to the commonly used clinical cutoff. Sociodemographic information, including age, place of residence, employment status, household income, and relationship status, was collected using a standardized questionnaire.

Statistical analysis. Continuous variables are presented as medians and interquartile ranges (IQR), and categorical variables as absolute and relative frequencies. Normality was assessed using the Shapiro-Wilk test. Because FSFI, number of missing teeth, OHIP-14, and age were non-normally distributed, Spearman rank correlation was used for bivariate analyses. FSFI scores according to relationship status were compared using the Mann-Whitney U test. Multivariable logistic regression was used to estimate the association of number of missing teeth and OHIP-14 score with sexual dysfunction (FSFI <=26.5), adjusted for age and relationship status. Results are expressed as odds ratios (OR) and 95% confidence intervals (CI). A two-sided p value <0.05 was considered statistically significant. Analyses were performed using GraphPad Prism 10.0 and Python (statsmodels 0.14). No formal a priori sample size calculation was performed; therefore, the analysis should be interpreted as exploratory.

## Results

Ninety-nine women were included. The median age was 42 years (IQR, 34-51). Most participants reported no professional activity (56.6%), and 23.2% reported a monthly household income below one minimum wage. Married women or women in a stable union represented 45.5% of the sample (Table 1).

**Table 1.**
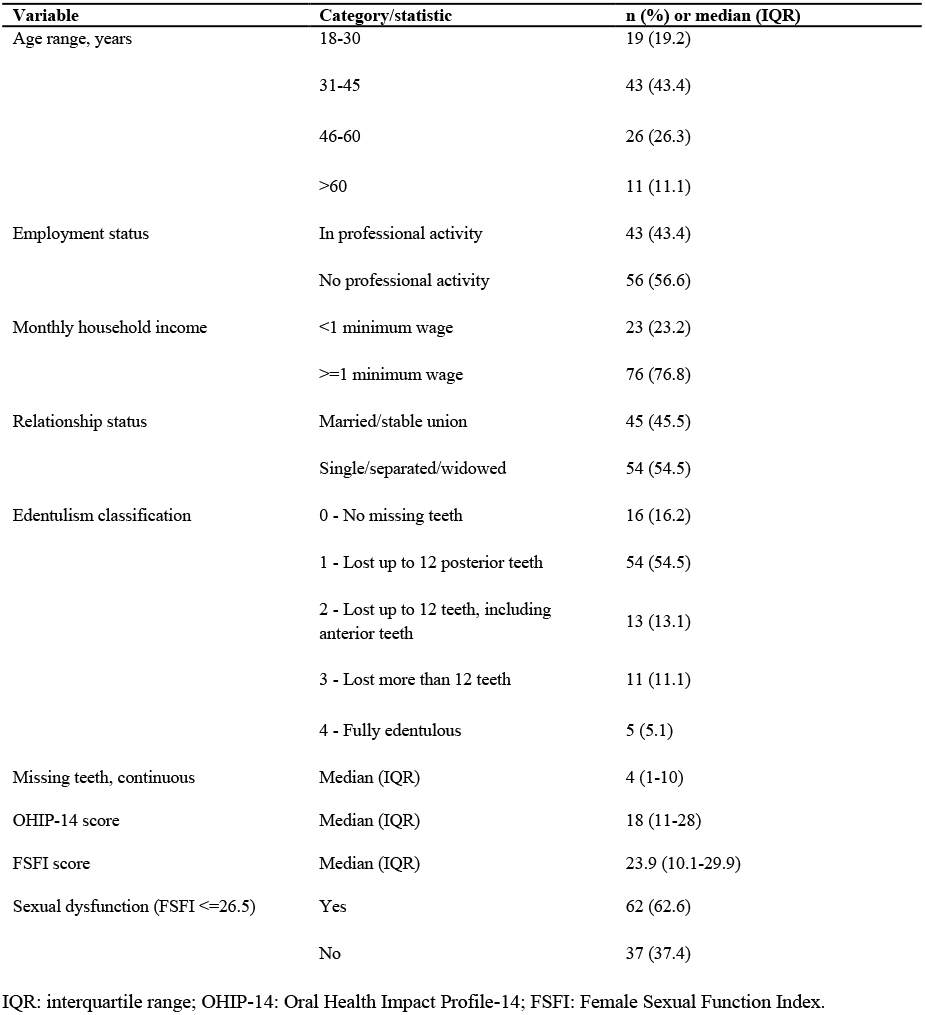
Sociodemographic, economic, and clinical characteristics of the study sample (N = 99).

Tooth loss was present in 83.8% of participants. The median number of missing teeth was 4 (IQR, 1-10). The median OHIP-14 score was 18 (IQR, 11-28), indicating a relevant burden of oral health impact. The median FSFI score was 23.9 (IQR, 10.1-29.9), and sexual dysfunction (FSFI <=26.5) was identified in 62.6% of women.

Spearman correlation analysis showed that FSFI scores were negatively associated with number of missing teeth (rho = −0.407; p < 0.001), OHIP-14 score (rho = −0.279; p = 0.005), and age (rho = −0.334; p < 0.001) (Figure 1). FSFI scores did not differ significantly between women married or in a stable union and the remaining participants (median, 24.6 vs. 18.6; Mann-Whitney U = 1,414; p = 0.162). In multivariable logistic regression, OHIP-14 score was independently associated with sexual dysfunction (OR = 1.05 per point; 95% CI, 1.01-1.10; p = 0.015). Number of missing teeth was not independently associated after adjustment (OR = 1.03 per additional missing tooth; 95% CI, 0.94-1.11; p = 0.536). Age and relationship status also did not reach statistical significance in the adjusted model (Table 2).

**Table 2.** Multivariable logistic regression for sexual dysfunction (FSFI <=26.5).

| Variable | OR (95% CI) | p value |
| --- | --- | --- |
| Missing teeth, per additional tooth | 1.03 (0.94-1.11) | 0.536 |
| OHIP-14 score, per point | 1.05 (1.01-1.10) | 0.015 |
| Age, per year | 1.03 (0.98-1.08) | 0.184 |
| Married/stable union | 0.85 (0.33-2.15) | 0.728 |

OR: odds ratio; CI: confidence interval; OHIP-14: Oral Health Impact Profile-14; FSFI: Female Sexual Function Index. The model included all variables listed in the table.

**Figure 1.**
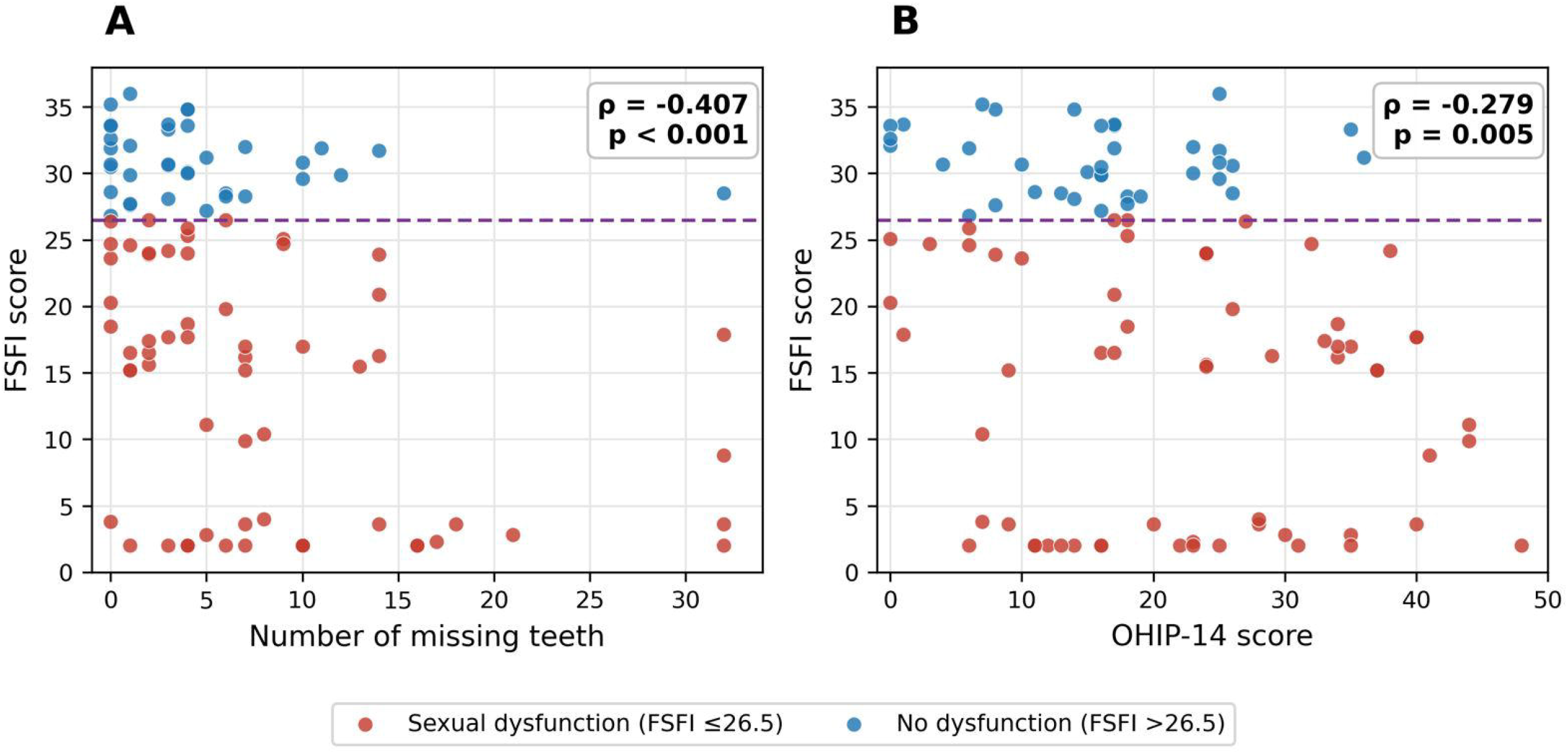
Scatterplots of FSFI total score according to (A) number of missing teeth and (B) OHIP-14 score. Red markers indicate participants with sexual dysfunction (FSFI <=26.5), and blue markers indicate participants without sexual dysfunction (FSFI >26.5). The dashed horizontal line indicates the FSFI cutoff for sexual dysfunction. No fitted regression line is shown because the bivariate analyses used Spearman rank correlation. FSFI: Female Sexual Function Index; OHIP-14: Oral Health Impact Profile-14.

## Discussion

This cross-sectional study found that tooth loss, worse OHRQoL, and older age were negatively correlated with FSFI scores in women attending primary dental care. In the adjusted logistic regression model, however, OHIP-14 score was the only independent factor associated with sexual dysfunction. The association between number of missing teeth and sexual dysfunction was attenuated after adjustment, suggesting that perceived oral health impact may be more closely related to sexual function than tooth count alone.

These findings should be interpreted cautiously. Because the study is cross-sectional and no formal mediation analysis was performed, the results do not demonstrate that OHRQoL mediates the association between tooth loss and sexual function. A more conservative interpretation is that the observed pattern is compatible with the hypothesis that OHRQoL partly explains or marks the pathway linking tooth loss to sexual well-being. Longitudinal studies are needed to determine whether tooth loss precedes changes in sexual function and whether improvements in OHRQoL after dental rehabilitation translate into improvements in FSFI scores.

The clinical relevance of these results lies in the broader effect of oral health on quality of life. Tooth loss may influence self-image, confidence, social interaction, and comfort in intimate relationships. Previous studies have shown that prosthetic rehabilitation can improve OHRQoL and aspects of sexual or social functioning, supporting the idea that dental care may have benefits beyond mastication and speech. In this context, OHIP-14 may capture psychosocial consequences of oral disease that are not fully represented by a simple count of missing teeth.

The study has strengths, including clinical assessment of tooth loss, use of standardized instruments, and adjustment for age and relationship status. It also has important limitations. The sample was relatively small and recruited from a single primary dental care service, limiting generalizability. Menopausal status, hormone therapy, psychiatric symptoms, medication use, active periodontal disease, prosthesis use, and partner-related factors were not systematically measured. The lack of an a priori sample size calculation also limits inferential strength. Future studies should include larger samples, validated measures of body image and mental health, detailed periodontal and prosthetic status, and longitudinal assessment before and after oral rehabilitation.

## Conclusion

Worse oral health-related quality of life was independently associated with sexual dysfunction in adult women attending primary dental care. Tooth loss was correlated with lower FSFI scores in bivariate analysis, but this association was attenuated after adjustment. These findings support the relevance of OHRQoL as an oral-health-centered marker associated with sexual well-being and reinforce the need to integrate oral rehabilitation and quality-of-life assessment into comprehensive women’s health care. Causal and mediational interpretations require confirmation in longitudinal studies.

## Data Availability

All data produced in the present study are available upon reasonable request to the authors.

